# A Single-Channel EEG Approach for Sleep Stage-Independent Automatic Detection of REM Sleep Behavior Disorder

**DOI:** 10.1101/2025.05.28.25328491

**Authors:** Gabriele Salvatore Giarrusso, Irene Rechichi, Alessandro Cicolin, Gabriella Olmo

## Abstract

Rapid Eye Movement (REM) Sleep Behavior Disorder (RBD) is a parasomnia characterized by the loss of physiological muscle atonia during REM sleep, often manifesting through dream-enacting behavior. Idiopathic RBD is largely considered a prodromal stage of neurodegenerative diseases, with a conversion rate to overt *α*-synucleinopathies of up to 96% after 14 years. Currently, the diagnostic procedure relies on time-consuming and labor-intensive inspection of polysomnography (PSG). This study proposes a Machine Learning (ML), stage-agnostic framework for the automatic detection of RBD subjects through unstaged, single-channel EEG sleep data from 58 subjects (32 RBD). The best model achieved 86.21% accuracy, 90.6% sensitivity, and 87.9% F-1 score, demonstrating strong predictive power. This study is the first to explore whole-night EEG data for RBD detection, paving the way for scalable, lightweight clinical decision support systems for early neurodegenerative screening and risk assessment.

**Clinical relevance:** This study presents a lightweight, clinical decision support tool to enhance RBD detection and support early interventions in neurodegenerative diseases.

## I. INTRODUCTION

Rapid Eye Movement (REM) Sleep Behavior Disorder (RBD) is a parasomnia characterized by the loss of physiological muscle atonia during REM sleep, which manifests through dream-enacting, oftentimes accompanied by violent behavior. Idiopathic RBD is considered a prodromal neurodegenerative stage, with a conversion rate to overt *α*-synucleinopathy of up to 96% after a 14-year follow-up [1]. Hence, this makes it a powerful tool as an early marker of neurodegenerative disorders such as Parkinson’s Disease (PD), Dementia with Lewy Bodies, and multiple system atrophy [2]. Criteria for the diagnosis of RBD require the lack of atonia, and the evidence of dream-enacting behavior [3], assessed through video-polysomnography (PSG) [4]. However, the process of scoring PSG, both for sleep staging and for subsequent diagnostic procedures, is manual and based on visual assessments by experts, resulting in a time-consuming, and labor-intensive procedure.

In the literature, various Machine Learning (ML) approaches have been proposed to support the diagnostic process of RBD [5], offering promising solutions through the automated analysis of PSG-derived signals – such as electroencephalography (EEG), electromyography (EMG), and electrooculography (EOG) [6], [7], [8], or inertial data [9]. Some advancements have been made by tackling Deep Learning (DL) approaches [10]. Recent studies specifically focused on RBD detection from EEG signals, particularly in the REM and Slow Wave Sleep (SWS) stages [11], [12], reporting encouraging results. However, a limitation persists: these approaches heavily rely on sleep staging, as they bank on hand-engineered features tailored to characterize the SWS and REM stages. To overcome this potential limitation, this study proposes a ML-based, stage-agnostic framework for the automatic detection of RBD subjects employing single-channel EEG sleep data. This solution not only promotes lightweight and minimally-invasive sleep studies but also seeks to simplify the diagnostic process by enhancing its robustness and scalability. The rest of the paper is structured as follows: **Section II** provides a description of the employed datasets, in terms of subjects and data. **Section III** illustrates the analysis pipeline, including signal pre-processing, feature extraction, and the ML framework. **Section IV** reports the experimental results and their interpretation; finally, **Section V** discusses the clinical implications of this work, addresses the limitations and highlights directions for future developments.

## II. MATERIALS

### A. Subjects

The study employed PSG recordings from 58 subjects – 26 Healthy Controls (HC) and 32 RBD subjects – from two sources: a public and a private datasets. The former is the CAP Sleep Database [13], publicly available on PhysioNet [14], consisting of PSG recordings of 16 HC (9 males, aged 32 *±* 5 years) and 22 RBD (19 males, aged 70 *±* 6 years), widely employed in the literature [6], [7]. The private database includes PSG data from 10 RBD subjects (8 males, aged 62 *±* 6 years) and 10 HC (6 males, aged 37 *±* 16 years). It was recorded at the Regional Center for Sleep Medicine (Molinette University Hospital, Turin) [7]; data collection was performed in concordance with the Declaration of Helsinki and approved by the local ethical committee (No. 00384/2020). The mean length across all recordings was 6.91 *±* 1.19 hours.

### B. Data

All PSG recordings were manually scored by sleep experts according to the American Academy of Sleep Medicine (AASM) scoring criteria [15], which classify sleep as REM sleep, and Non REM (NREM) sleep (further divided in N1, N2, SWS, from light to deep). As previously introduced, and in the perspective of developing lightweight screening tools for supporting sleep studies, this work relies on a single EEG channel and adopts only essential pre-processing steps. In more detail, only recordings from the central EEG channel (C3-A2, or C4-A1) were considered. Then, EEG recordings were exploited and merged, so as to obtain two different feature sets to be employed in the ML framework. Specifically, for the first feature set, the whole NREM sleep (stages SWS, N2, N1) was considered as a uniform segment. Then, for the second feature set, a comprehensive approach was tackled by taking into account the whole sleep, regardless of the stage (REM, SWS, N2, N1). This procedure is further detailed in Section III-A.

## III. METHODS

This section describes the procedures adopted in the analysis, carried out through custom-written code in MATLAB® vR2023b and Python v3.11.

### A. Signal Pre-Processing

The signals underwent minimal pre-processing, to ensure applicability in natural environments and wearable scenarios. The EEG recordings were band-pass filtered (0.01–40 Hz) through a cascade of two zero-phase, anti-causal Butterworth filters – low-pass and high-pass. Then, the EEG segments were processed so as to follow two different analysis pipelines (Figure 1). Specifically, as described earlier, first only NREM segments were considered (SWS, N2, N1), then, all sleep was taken as a whole, stage-independent segment. Finally, features were extracted separately on the two EEG segments, as described in the following section.

**Fig. 1.**
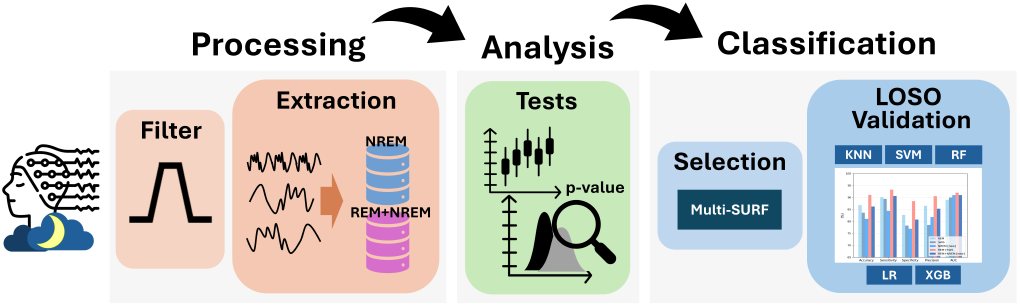
Pipeline chart.

### B. Feature Extraction

A comprehensive, multi-domain extraction process was carried out, considering key attributes found in topic-related studies [16], [17], [18], [11], [8], along with the division of EEG features into Time Domain (TD), Frequency Domain (FD), Time-Frequency Domain (TFD), and Nonlinear Domain (NND). This guaranteed the inclusion of morphological and spectral characteristics of EEG, together with details regarding its non-stationary and non-linear nature, expression of the underlying intricate brain mechanisms from which it derives. Table I summarizes the employed features; they were extracted in 30-seconds epochs, to match the AASM sleep scoring criteria. To ensure EEG signal stationarity, 2-seconds windows were employed for the FD, TFD, and NND features [12]. Several statistics, including Mean, Standard Deviation, 75^*th*^ percentile, and Main Mode (MM) were then employed to summarize the obtained values, as in [12], and employed as distinct features (Figure 2). This procedure, resulting in a (58, 900) feature matrix, was repeated for all patients, eventually obtaining two feature sets:

**TABLE I.** Features employed in the study, according to their domain. *⋄*: adapted from cited study.

| Feature and Domain | Reference |
| --- | --- |
| *Time Domain* |  |
| Mean, mode, STD, skewness, kurtosis, maximum and minimum values, Range | ◊[19] |
| Zero Crossing Rate | ◊[20], [11] |
| Hjorth Parameters (activity, mobility, complexity) | ◊[11] |
| Hjorth-based parameters (sparseness, irregularity, spectrum bandwidth, average-zero-up, crossing period, average-peak-to-peak period) | ◊[12] |
| Percentiles (25, 75), interquartile range | ◊[19] |
| Form, Crest, and Impact factors | [11] |
| Coastline | [12] |
| *Frequency Domain* |  |
| Absolute Power Spectrum, Mean Power Spectrum | ◊[19] |
| Peak Frequency, Mean Frequency (Spectral Centroid), Median Frequency (SEF50), SEF25, SEF75, SEF95, SEFd75-25, SEFd95-25, SEFd95-50 | ◊[19], [11], [12] |
| Spectral Crest, Spectral Entropy, Renyi's quadratic Entropy | [20] |
| *Frequency (EEG sub-bands)* |  |
| Max Value, Relative Power Spectrum, Mean Power Spectrum, Absolute Power Spectrum | ◊[19], [12] |
| Spectral Centroid, Spectral Spread, Spectral Skewness, Spectral Kurtosis, Variance of Central Frequency | ◊[12] |
| Peak Frequency, Median Frequency (SEF50), SEF95, SEFd95-50 | ◊[11] |
| Bandpower-ratios: $\theta/\alpha$, $\beta/\alpha$, $(\theta+\alpha)/\beta$, $\theta/\beta$, $(\theta+\alpha)/(\alpha+\beta)$ | ◊[20] |
| TREM/FREM, SWA/SOs | ◊[19], [11], [12] |
| Spectral Entropy, Renyi's quadratic Entropy | ◊[20] |
| *Time-Frequency* |  |
| 6 levels Discrete Wavelet Analysis (DWT), db4 (Mean, Standard Deviation, Coastline, Ratio of absolute mean values of adjacent sub-bands) | ◊[19] |
| *Non Linear* |  |
| Detrended Fluctuation Analysis | [20] |
| Higuchi Fractal Dimension | ◊[19] |
| Sample Entropy, Taiger-Kaiser Energy Operator (Mean, Standard Deviation, Skewness, Kurtosis, Maximum value) | ◊[20], [12] |

**Fig. 2.**
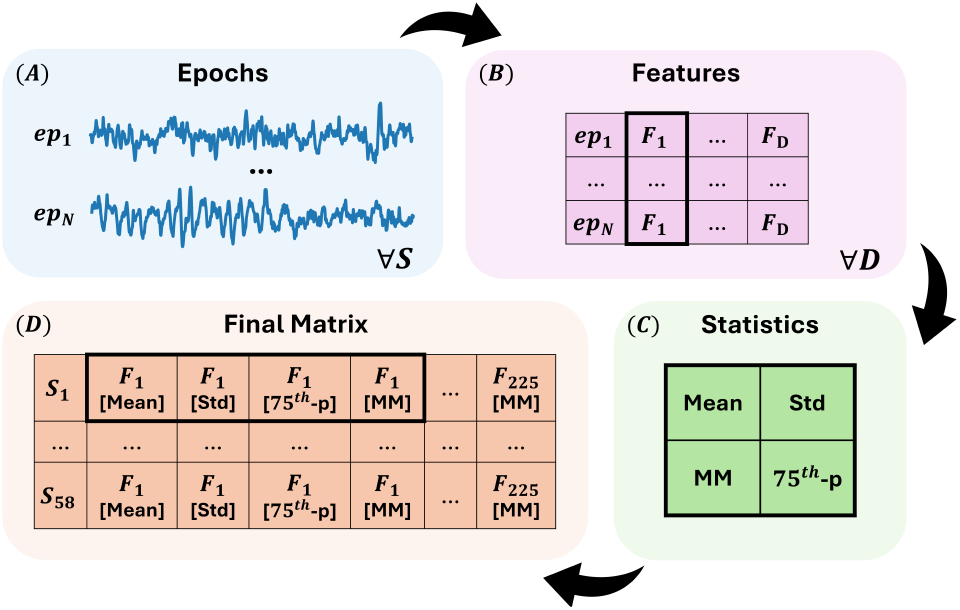
**(A)** Each EEG signal is divided into 30-seconds (TD) or 2-seconds (FD, TFD, NND) epochs, based on the specific domain D. **(B)** Features F (TD: 32, FD: 165, TFD: 20, NND: 8, total: 225) are extracted for each epoch, and then **(C)** summarized through four statistics, namely Mean, Standard Deviation, 75^*th*^ percentile, and Main Mode, resulting in 900 features. The process is repeated for each Subject S (1,…, 58), **(D)**, resulting in a feature matrix (58, 900).

- *Set*_*NREM*_ : features extracted from (*SWS* + *N* 2 + *N* 1) sleep
- *Set*_*ALL*_: features extracted from (*REM* + *SWS* + *N* 2 + *N* 1) sleep

### C. Statistical Analysis, Feature Normalisation, and Feature Selection

Statistical tests were performed on the extracted features to test their relevance for the task, through the open-source software Jamovi [21]. First, a Shapiro-Wilk normality test was carried out; then, independent samples testing was conducted, through the Student’s t-test and Mann-Whitney U test for normally and non-normally distributed features, respectively. Z-score normalization was performed on the analyzed attributes. Feature selection was conducted in both feature sets through the Multiple Threshold Spatially Uniform ReliefF (MultiSURF) feature selection method [22]. It is a ReliefF-based algorithm [23], which proved to be effective in highdimensional biomedical datasets, possibly with non-linear relations among features, as the ones explored in this study. Features were ranked according to their importance score, and the top-10 features were employed for the classification task. For the sake of clarity, feature selection was performed in all training folds to limit the risk of data leakage.

### D. Classification Framework

Supervised ML models were employed to carry out the automatic detection of RBD subjects. Namely, a Support Vector Machine (SVM), a K-Nearest Neighbors (KNN), a Random Forest (RF), an eXtreme Gradient Boosting (XGBoost), and a Logistic Regression (LR) classifier were adopted for both feature sets. Given the relatively small sample size (58 total subjects, 38 RBD), a Leave-One-Subject-Out (LOSO) Cross Validation (CV) approach was adopted to ensure proper generalization of the models. In this procedure, model training is repeated *N* times (*N* : total number of subjects); at each iteration, the classifier is trained on (*N −* 1) subjects, and tested on the held-out subject. Hyperparameters were optimized on training data only, through a *k*-fold (*k*=5) combined with a Grid Search approach, to further enhance the robustness of the models. Table II provides a summary of the employed classifiers and the optimized parameters, along with their search range.

**TABLE II.** Summary of the employed classifiers and the searched hyperparameters, (parameter and range).

| Model | Searched Hyperparameters |
| --- | --- |
| KNN | *Number of neighbors (K)*: [1–20]<br>*Distance*: Minkowski, Euclidean, Manhattan, Chebyshev |
| SVM | *Kernel function*: polynomial, radial basis, sigmoid<br>$\gamma$: [0.0001, 0.001, 0.01, 0.1, 1]<br>*Penalty (C)*: [0.01, 0.1, 1, 10, 100, 1000] |
| RF | *Number of trees*: [10, 50, 100, 150]<br>*Criterion*: gini, entropy, logarithmic loss<br>*Max Depth*: [1, 2, 3, 5]<br>*Max features*: sqrt, log2 |
| XGBoost | *Learning rate*: [0.001, 0.01, 0.1, 1]<br>*Gamma*: [0.0001, 0.001, 0.01, 0.1]<br>*Max Depth*: [1, 3]<br>*Number of trees*: [25, 50, 100] |
| LR | *Penalty (C)*: [0.001, 0.01, 0.05, 0.1, 1, 10, 50, 100]<br>*Max iterations*: [15, 20, 25, 50]<br>*Solver*: Newton-CG, linear |

## IV. RESULTS

### A. Feature Inspection

As mentioned in Section III-B, a total of 225 features were extracted divided according to the domain. Specifically: 32 for TD, 165 for FD, 20 for TFD, and for in NDD (Figure 2); for each feature, a total of four statistics were computed and employed as predictors, resulting in the (58, 900) final feature set (58 subjects, cf. III-B). The independent samples testing revealed a substantially large number of features exhibiting high-to-very-high statistical significance (*p* :*<* 0.001 *−* 0.05). In an attempt to limit the computational load required in the classification step, a preliminary data reduction step was carried out by keeping only features with *p <* 0.05 (Mann-Whitney U). This resulted in *Set*_*NREM*_ set to have 361 features and 347 for *Set*_*ALL*_. Feature selection was repeated on all training folds; as the MultiSURF approach ranks features according to their importance, the resulting top-10 features in each fold were used for classification. Table III reports the top-10 most recurrent features across all training folds, in terms of Frequency of Occurrence (FO) (%). As appreciable, PKF_*FREM*_, representing the peak frequency in the FREM band (2–8 Hz), proved to be a meaningful predictor for detecting RBD, being selected in 98% and 100% of folds in the *Set*_*NREM*_ (PKF_*FREM,Mean*_) and *Set*_*ALL*_ sets (PKF_*FREM,Std*_), respectively. The most recurrent feature in *Set*_*NREM*_ was SEFd(95-50)_*θ,Mean*_ (100% of folds), which appeared significantly predictive also in *Set*_*ALL*_ (95% of folds). Considering only feature importance, in the *Set*_*NREM*_ set, the most informative feature was SSp_*β,Std*_ (STD of the spectral skewness in *β*-band), ranked first in 79% of folds. This parameter is also featured among the most recurrent features, being selected overall in 98% of all training folds. Regarding *Set*_*ALL*_, the feature PKF_*FREM,Std*_, selected in the totality of training folds, emerged also as the most informative for the classification, being ranked top position in 69% of folds. These results suggest that the *β* EEG band holds great predictive power for detecting RBD through sleep EEG, as also supported by [24]. Similar considerations can be done for FREM (2–8 Hz), in line with [18], and the *θ* band. Figure 3 and Figure 4 depict the distribution (obtained through Kernel Density Estimation) in the two classes of the most recurrent features for *Set*_*NREM*_ and *Set*_*ALL*_, respectively. As appreciable, these plots further confirm the discriminative power of the selected parameters.

**TABLE III.**
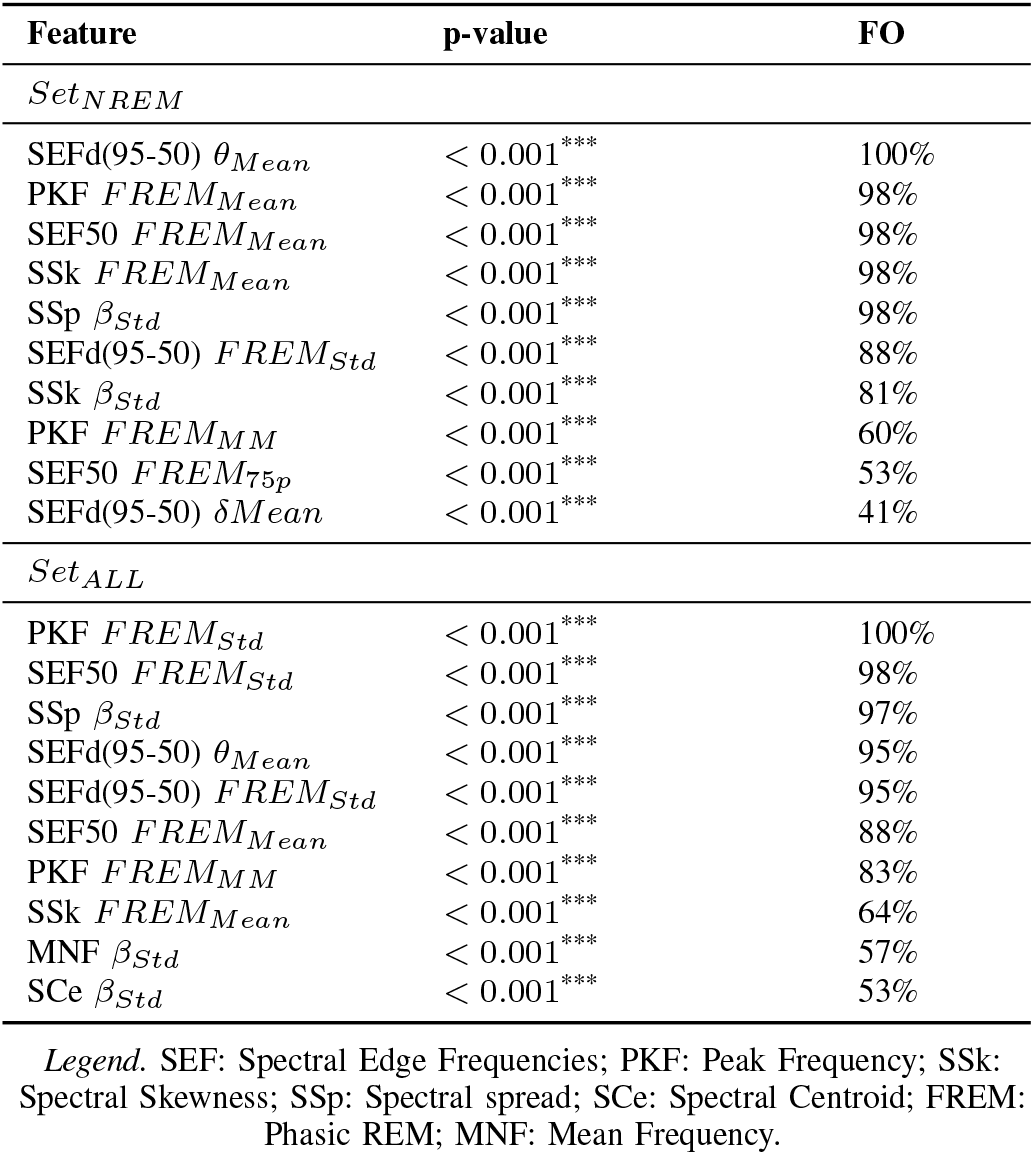
Top 10 most recurrent Features in the two training sets, with frequency of occurrence (FO) and statistical significance ^***^ : *p <* 0.001. *<* 0.001^***^ 53%.

| Feature | p-value | FO |
| --- | --- | --- |
| *Set<sub>NREM</sub>* |  |  |
| SEFd(95-50) $\theta_{Mean}$ | $< 0.001^{***}$ | 100% |
| PKF $FREM_{Mean}$ | $< 0.001^{***}$ | 98% |
| SEF50 $FREM_{Mean}$ | $< 0.001^{***}$ | 98% |
| SSk $FREM_{Mean}$ | $< 0.001^{***}$ | 98% |
| SSp $\beta_{Std}$ | $< 0.001^{***}$ | 98% |
| SEFd(95-50) $FREM_{Std}$ | $< 0.001^{***}$ | 88% |
| SSk $\beta_{Std}$ | $< 0.001^{***}$ | 81% |
| PKF $FREM_{MM}$ | $< 0.001^{***}$ | 60% |
| SEF50 $FREM_{75p}$ | $< 0.001^{***}$ | 53% |
| SEFd(95-50) $\delta_{Mean}$ | $< 0.001^{***}$ | 41% |
| *Set<sub>ALL</sub>* |  |  |
| PKF $FREM_{Std}$ | $< 0.001^{***}$ | 100% |
| SEF50 $FREM_{Std}$ | $< 0.001^{***}$ | 98% |
| SSp $\beta_{Std}$ | $< 0.001^{***}$ | 97% |
| SEFd(95-50) $\theta_{Mean}$ | $< 0.001^{***}$ | 95% |
| SEFd(95-50) $FREM_{Std}$ | $< 0.001^{***}$ | 95% |
| SEF50 $FREM_{Mean}$ | $< 0.001^{***}$ | 88% |
| PKF $FREM_{MM}$ | $< 0.001^{***}$ | 83% |
| SSk $FREM_{Mean}$ | $< 0.001^{***}$ | 64% |
| MNF $\beta_{Std}$ | $< 0.001^{***}$ | 57% |
| SCe $\beta_{Std}$ | $< 0.001^{***}$ | 53% |

Legend. SEF: Spectral Edge Frequencies; PKF: Peak Frequency; SSK: Spectral Skewness; SSP: Spectral spread; SCe: Spectral Centroid; FREM: Phasic REM; MNF: Mean Frequency.

**Fig. 3.**
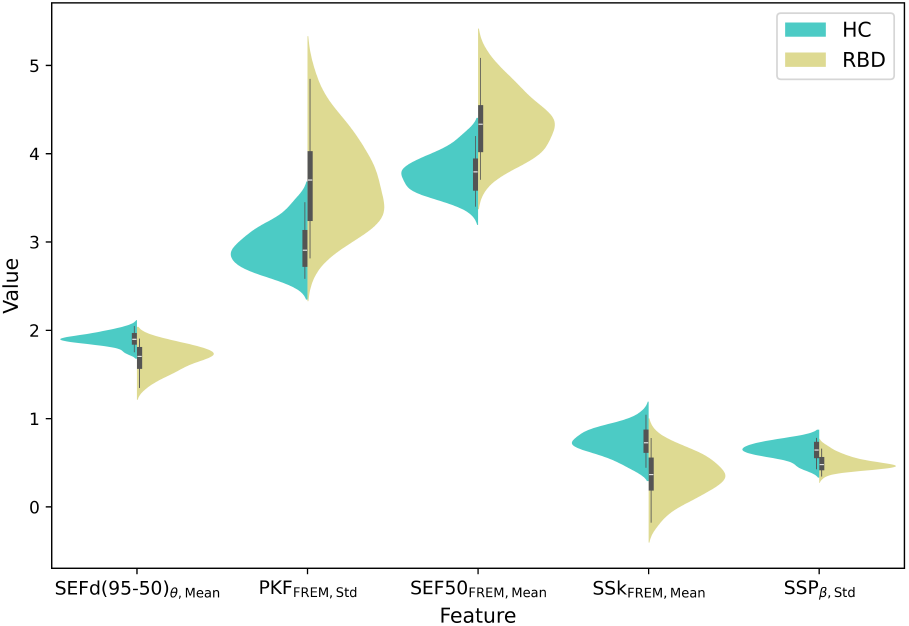
Distribution of the top 5 most recurrent features in *Set*_*NMREM*_, ordered following descending FO values.

**Fig. 4.**
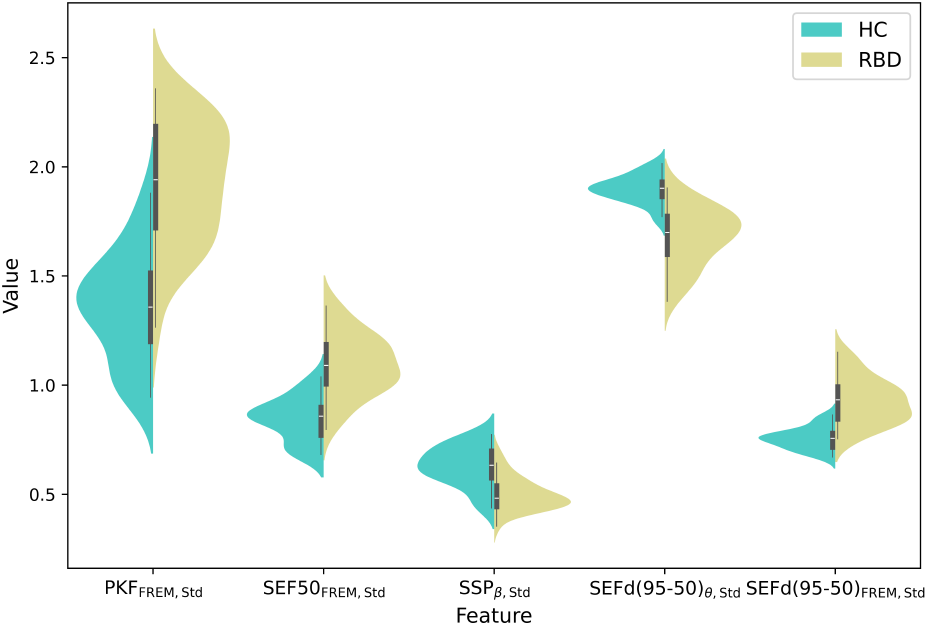
Distribution of the top 5 most recurrent features in *Set*_*ALL*_, ordered following descending FO values.

### B. RBD Detection Performance

The explored classifiers exhibited moderate-to-high classification accuracies in both feature sets, achieving Macro Averaged Accuracy (MAA) values of 78.28 *±* 0.02 % and 81.03 *±* 0.03 % for *Set*_*NREM*_ and *Set*_*ALL*_, respectively, across all models. These results not only indicate robust classification performance but also suggest the high predictive power of the selected parameters, as already revealed in the statistical analysis. Notably, these results become significant considering the minimal signal processing applied to the EEG, suggesting the practicability of the proposed approach. The Area Under the Curve (AUC) values, across the models, of 0.89 *±* 0.02 for *Set*_*ALL*_ and 0.86 *±* 0.03 for *Set*_*NREM*_ further demonstrate the strong discrimination ability for the two classes (HC and RBD), which remains consistent through the two feature sets. For the sake of brevity, Table IV reports the LOSO-CV classification performance of the best models in the two feature sets, in terms of Accuracy, Sensitivity, Precision, F1-score, and AUC. In *Set*_*NREM*_ a LR (C=50, max iterations: 15, solver: Newton-CG) emerged as the best model, with 81% Accuracy, 84.4% Sensitivity, and an AUC of 0.91. As appreciable, higher performance metrics were attained in *Set*_*ALL*_; the best model was an XGBoost (eta: 0.1, gamma: 0.0001, max depth: 3, number of trees: 25), showing an 86.2% Accuracy, 90.6% Sensitivity, and 0.91 AUC, suggesting high predictive power of the selected features. A moderately high value of F1-score was also obtained (87.9%), indicative of good model robustness. These performances are in line with previous works employing the same set of subjects [7], [12], which achieved a MAA of 86.21%, and AUC of 0.92 (Figure 6). Although seemingly lower than those obtained through stage-specific approaches (Figure 5), the performance of this study are encouraging, particularly taken into account that continuous sleep data exhibit very diverse characteristics. As shown in Figure 5, *Set*_*NREM*_ yields lower Accuracy, Sensitivity, and Specificity compared to SWS alone, but exhibits higher Precision and AUC. This suggests that *Set*_*NREM*_ is less prone to false positives while maintaining a good detection trade-off across various thresholds. Likely, the presence of lighter and more variable sleep stages (N1, N2) could possibly introduce confounding elements compared to the more stable SWS. Nevertheless, a direct comparison with previous works is challenging, as the analyses in [7], [12] relied on hand-engineered features from specific and distinct sleep stages – i.e., SWS and REM. To the best of the Authors’ knowledge, this study is the first to consider the whole EEG sleep segments without stage-dependent feature extraction, making the analysis more generalizable and clinically practical. Additionally, as displayed by the confusion matrix in Figure 7, the relatively low number of False Negatives (FN) in *Set*_*ALL*_ further supports the feasibility of the proposed approach, suggesting its suitability to clinical screening applications, that seek to minimize the number of FN.

**TABLE IV.** RBD classification performance (%) of the best algorithms.

|  | *Set<sub>NREM</sub>* | *Set<sub>ALL</sub>* | *Benchmark* |
| --- | --- | --- | --- |
| Accuracy | 81.03 | 86.21 | 91.09 ± 0.00 |
| Sensitivity | 84.38 | 90.63 | 93.23 ± 0.03 |
| Specificity | 76.92 | 80.77 | 88.47 ± 0.04 |
| Precision | 81.82 | 85.29 | 90.55 ± 0.02 |
| F1 | 83.08 | 87.88 | 92.03 ± 0.00 |
| AUC | 0.91 | 0.91 | 0.92 ± 0.01 |
| MAA | 78.28 ± 0.02 | 81.03 ± 0.03 | 86.21 ± 0.03 |

Best model for *Set<sub>NREM</sub>*: LR; Best model for *Set<sub>ALL</sub>*: XGBoost. The third column represents the averaged results obtained using merging REM+SWS data in [11], [12], used here as a benchmark of best results.

**Fig. 5.**
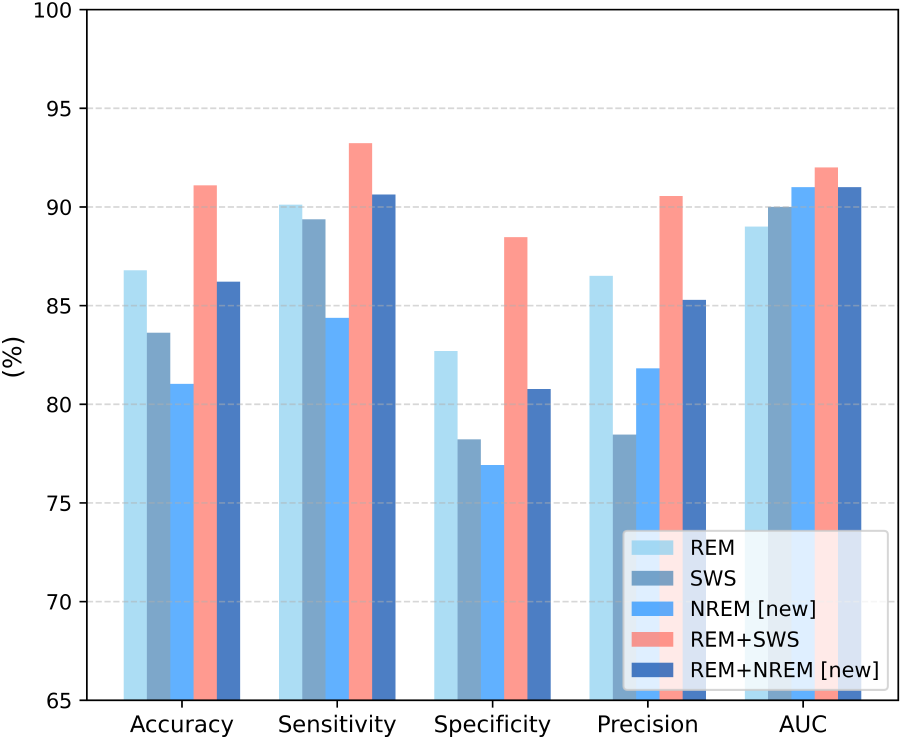
Performance comparison with topic-related studies. SWS, REM, and REM+SWS represent the averaged results obtained using various REM and SWS data combinations in [11], [12]. NREM and REM+NREM represent the novelty of the current work.

**Fig. 6.**
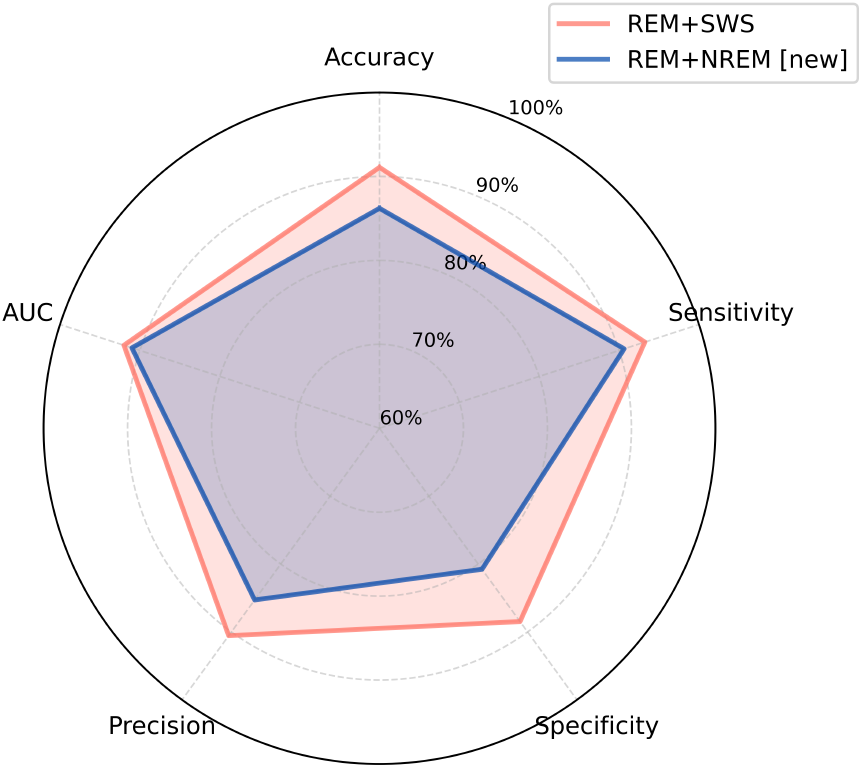
Performance comparison with best results in previous works. For the sake of clarity: (REM+SWS) represents the averaged results from [11], [12], (REM+NREM) represents the novelty of the current work.

**Fig. 7.**
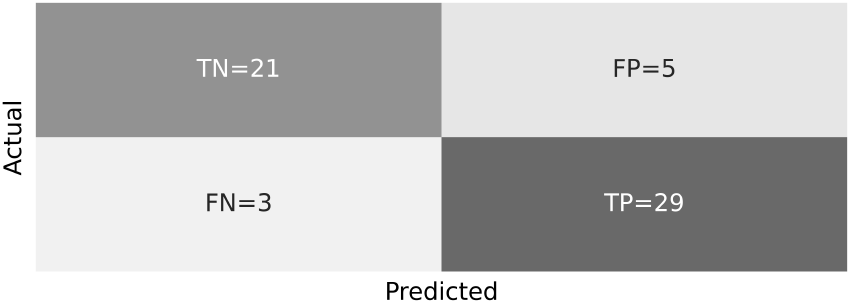
Confusion matrix for the best model in *Set*_*ALL*_ (XGBoost).

## V. CONCLUSION AND FUTURE WORK

RBD is largely considered a prodrome to *α*-synucleinopathies, making it an important actor in neurodegenerative diseases. Currently, its diagnosis requires protracted manual labor, significantly impacting diagnostic times, and possibly leading to interrater variability. Although various ML have been proposed to aid the diagnostic process, they still rely on sleep staging (either manual or automated), inevitably requiring full PSG assessments, with significant patient discomfort.

In the perspective of developing minimally-invasive, scalable sleep studies, this paper proposes a ML-based framework for the automatic detection of RBD subjects, through the analysis of single-channel EEG and stage-agnostic features. The preliminary investigation results obtained in *Set*_*NREM*_, when compared with the previous works from the same research group [11], [12], suggest that sleep stages N1 and N2 have a minimal contribution in the binary classification task, fostering the feasibility and practicability of a stage-agnostic approach. Indeed, the performance metrics in *Set*_*ALL*_ (86.21% Accuracy, and Sensitivity above 90%), although slightly lower compared to stage-specific approaches on the same datasets [11], [12], suggest the feasibility of efficiently detecting RBD through unstaged EEG. Indeed, a Cohen’s *κ* of 72% was obtained with the best classification algorithm (XGBoost) on *Set*_*ALL*_, indicative of substantial agreement between the developed framework and manual assessment. Hence, these findings support the development of clinical decision support systems for RBD without requiring a full-PSG evaluation and conventional sleep scoring, to be used in large-scale screening protocols for neurodegenerative risk assessment.

Although with promising results, this study is not without limitations. First, this framework should be tested on a larger number of subjects, possibly well-stratified for sex and age, in order to properly assess its scalability and applicability. Second, due to computational load restrictions, the Grid Search for hyperparameters optimization was conducted on a limited combination of parameters ranges.

Future implementations should test more extensive parameters ranges with possibly beneficial effects on the classification performance. Moving forward, the best-performing model derived from *Set*_*ALL*_ (cf. Section IV-B) and the top-10 most recurrent features (illustrated in Table III), will be employed on unseen data to assess the proposed framework’s robustness and to externally validate its predictive capabilities. Finally, further tests with EEG recorded through wearable technology should be tackled, to assess the robustness and reliability of the proposed systems in natural environments.

Overall, this study demonstrates the feasibility of employing unstaged EEG for the automatic detection of RBD, supporting advancements in lightweight, scalable clinical decision support systems for early identification of neurodegenerative states.

## Data Availability

All data produced in the present study are available upon reasonable request to the authors

## ACKNOWLEDGMENT

The drafting and publication of this work took place for G.G. in the context of the project PNRR-NGEU which has received funding from the MUR-DM 117/2023 (Italy), and for I.R. of the project PNRR-NODES which has received funding from the MUR-M4C2 1.5 funded by the European Union NextGenerationEU (Grant agreement no. ECS00000036).

